# Risk factors for severe corona virus disease 2019 (COVID-19) patients : a systematic review and meta analysis

**DOI:** 10.1101/2020.03.30.20047415

**Authors:** Lizhen Xu, Yaqian Mao, Gang Chen

**Author notes:** Correspondence to: Gang Chen, Fujian Academy of Medical Sciences, Fuzhou 350001, China. The first two authors contributed to the study equally.

## Abstract

**Importance:** With the increasing number of infections for COVID-19, the global health resources are deficient. At present, we don’t have specific medicines or vaccines against novel coronavirus pneumonia (NCP) and our assessment of risk factors for patients with severe pneumonia was limited. In order to maximize the use of limited medical resources, we should distinguish between mild and severe patients as early as possible.

**Objective:** To systematically review the evidence of risk factors for severe corona virus disease 2019 (COVID-19) patients.

**Evidence Review:** We conducted a comprehensive search for primary literature in both Chinese and English electronic bibliographic data bases including China National Knowledge Infrastructure (CNKI), Wanfang, Weipu, Chinese Biomedicine Literature Database (CBM-SinoMed), MEDLINE (via PubMed), EMBASE, Cochrane Central Register, and Web of science. The American agency for health research and quality (AHRQ) tool were used for assessing risk of bias. Mata-analysis was undertaken using STATA version 15.0.

**Results:** 20 articles (N=4062 participants) were eligible for this systematic review and meta-analysis. First in this review and meta-analysis, we found that elderly male patients with a high body mass index, high breathing rate and a combination of underlying diseases (such as hypertension, diabetes, cardiovascular disease, and chronic obstructive pulmonary disease) were more likely to develop into critically ill patients. second, compared with ordinary patients, severe patients had more significant symptom such as fever and dyspnea. Besides, the laboratory test results of severe patients had more abnormal than non-severe patients, such as the elevated levels of white-cell counts, liver enzymes, lactate dehydrogenase, creatine kinase, c-reactive protein and procalcitonin, etc, while the decreased levels of lymphocytes and albumin, etc.

**Interpretation:** This is the first systematic review investigating the risk factors for severe corona virus disease 2019 (COVID-19) patients. The findings are presented and discussed by different clinical characteristics. Therefore, our review may provide guidance for clinical decision-making and optimizes resource allocation.

**Key Points:** *Question:* What are the risk factors for severe patients with corona virus disease 2019 (COVID-19)?

*Findings:* First in this review and meta-analysis, we found that elderly male patients with a high body mass index, high breathing rate and a combination of underlying diseases were more likely to develop into critically ill patients. second, compared with ordinary patients, severe patients had more significant symptom such as fever and dyspnea. Last, we also found that the laboratory test results of severe patients had more abnormal than non-severe patients.

*Meaning:* This review summaried the risk factors of severe COVID-19 patients and aim to provide a basis for early identification of severe patients by clinicians.

## Introduction

In December 2019, COVID-19 was discovered in Wuhan, Hubei Province, China^[1]^. On January 7, 2020, the causative agent was identified as a 2019 novel coronavirus (2019-nCoV)^[2-4]^. According to the WHO report^[5]^, as of 10:00 on March 23, 2020, a total of 334,695 confirmed cases and 14,526 deaths were reported in 140 countries and regions worldwide. While the epidemic in China is gradually under control, Europe and the Middle East have shown the signs of rapid spread. For example, Italy, Iran, Germany, Spain, the United States, South Korea and Japan have seen a sharp increase in the number of infections^[5]^.

Nowadays, with the increasing number of infections, the global health resources are extremely poor. In order to maximize the use of limited medical resources, we should distinguish between mild and severe patients as early as possible. At present, we don’t have specific medicines or vaccines against novel coronavirus pneumonia (NCP) and our assessment of risk factors for patients with severe pneumonia was limited. In this regard, we have summarized the published studies with critically ill patients aimed at explaining the risk factors of the NCP and providing Chinese experience for people around the world in responding to COVID-19.

## Methods

This meta-analysis was performed in accordance with PRISMA-2009 (Preferred Reporting Items for Systematic Reviews and Meta-analyses)^[6]^ and MOOSE (Meta-analysis of Observational Studies in Epidemiology) guidelines^[7]^.

### Search strategy

Relevant studies were searched from both Chinese and English electronic bibliographic databases including China National Knowledge Infrastructure (CNKI), Wanfang, Weipu, Chinese Biomedicine Literature Database (CBM-SinoMed), MEDLINE (via PubMed), EMBASE, Cochrane Central Register and Web of science from inception to 8 March 2020. The MeSH terms of COVID-19 and corresponding synonyms were included into the searching strategy. Limiting our search to human-subjects but without language limitation. Reference lists of retrieved articles were also reviewed to further identify potentially relevant studies. The searching procedures were independently performed by two reviewers (Lizhen Xu and Yaqian Mao) and disagreements were settled by discussion.

### Inclusion criteria

The inclusion criteria were as follows: (i) prospective or retrospective original literature; (ii) all patients were clearly diagnosed with COVID-19; (iii) characteristics of severe and non-severe patients were documented; and (iv) complete medical records were available for data extraction.

Definition of severe COVID-19^[8]^: Severe COVID-19 was designated when the patients had one of the following criteria:1) Respiratory distress with respiratory frequency≥30/min; 2) Pulse Oximeter Oxygen Saturation≤93% at rest; 3) Oxygenation index (artery partial pressure of oxygen/inspired oxygen fraction, PaO2/FiO2)≤300 mmHg (1 mmHg=0.133 kPa). At high altitudes (above 1000 meters), PaO2 / FiO2 should be corrected according to the following criteria: PaO2 / FiO2 [Atmospheric Pressure (mmHg) / 760]. Pulmonary imaging showed that the lesions progressed more than 50% within 24-48 hours should be managed according to Critically ill patients.

### Quality assessment

The observational study quality evaluation criteria recommended by the American agency for health research and quality (AHRQ) were used to evaluate the study quality. These criteria included 11 items, including subjects selection, research quality control and data processing. Each question will be answered with “yes,” “no” or “unclear.”

### Data extraction

The following characteristics were extracted from selected studies: authors, sample size, region, study period. In addition, the following possible risk factors were recorded independently: patient characteristics, comorbidities, vital signs, symptoms, laboratory findings. Data extraction was accomplished independently by two reviewers (Lizhen Xu and Yaqian Mao). Any disagreement was resolved by joint discussion to reach a consensual conclusion. There were a few points that need to be explained. First, in order to minimize the risk of duplication of data, when two or more studies presented possible overlap, where sampling periods overlapped and patients were from the same region, the one with largest populations was included. Second, continuous variables were expressed as medians and interquartile ranges or simple ranges in some studies, the standard deviation and mean value were not estimated due to inaccurate.

### Data synthesis and statistical analysis

For categorical variables, analysis was performed by calculating the odds ratio (OR) with 95% confidence interval (95% CI). For continuous outcomes, Weighted Mean Difference (WMD) and standardized mean difference (SMD) with the corresponding 95% CI were calculated. The heterogeneity was assessed by using the *I*^*2*^ test, with *I*^*2*^>50 % indicating the existence of heterogeneity. If there is significant heterogeneity, a random effect model (DerSimonian-Laird method) was used to calculate the pooled effect; otherwise, the fixed model (Mantel-Haenszel method) was used. Possible publication bias was evaluated via observing the symmetry characteristics of funnel-plots. If the number of included studies in each outcome was <10, the funnel plot was not performed due to limited power^[9]^. Data analysis was undertaken using STATA, version 15.0 (Stata Corp).

## Results

### Literature search results

The entire process of literature searching and screening was displayed in **Figure 1**. Initially, 6354 publications were identified through database searching. After the removal of 2622 duplicates, there remained a record of 3732 studies. We excluded 3654 records by reviewing their titles and abstracts. As a result, only 78 articles were subject to a full-text review. Finally, 20 articles met the inclusion criteria were included in the synthesis.

**Figure 1.**
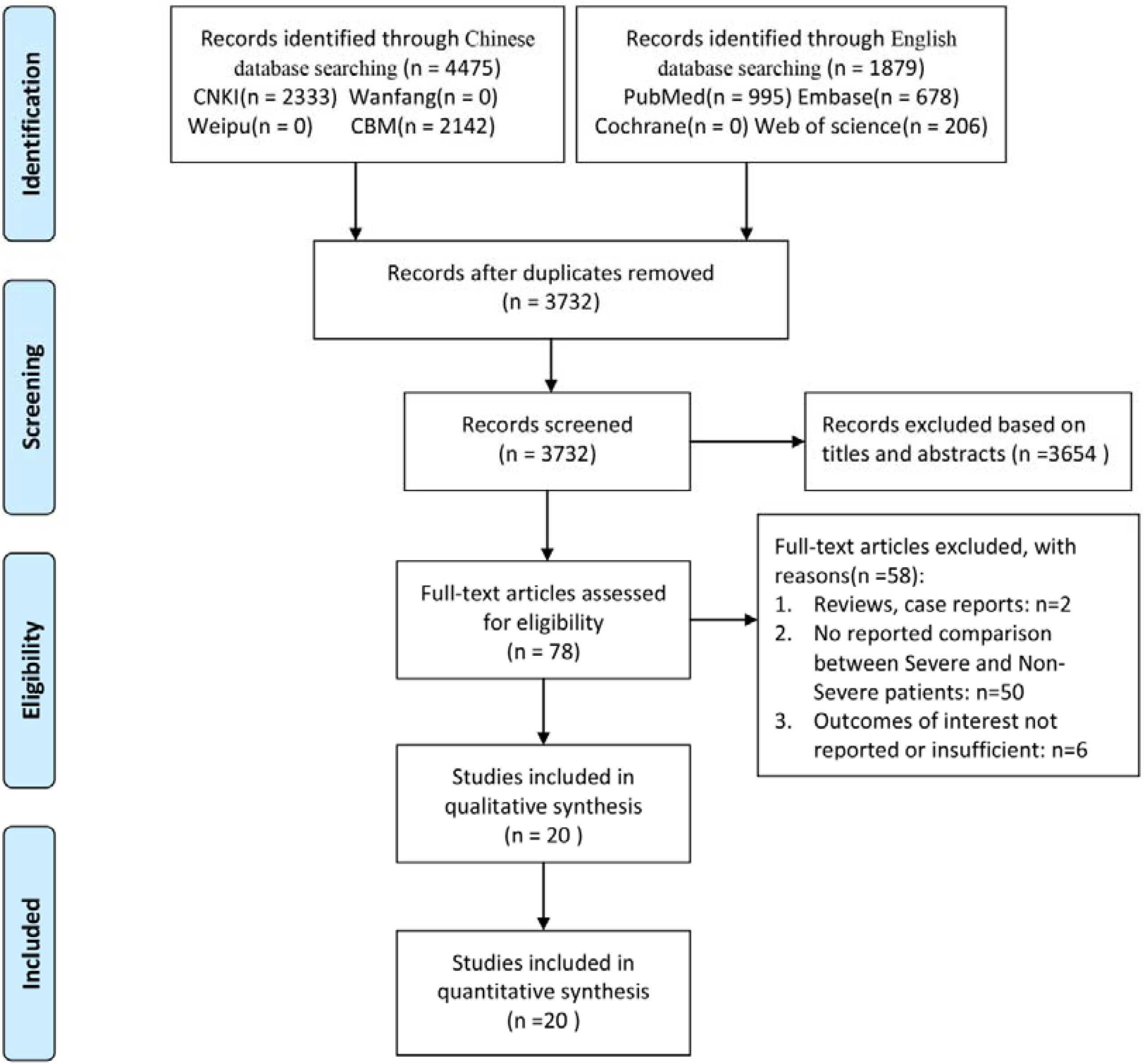
A schematic flow diagram of studies’ search and retrieval process.

### Study characteristics

The main characteristics of the studies were summarized in **Table 1**. All articles were observational studies. All the subjects are from China, including more than 30 provinces and cities. The study period spans from 11 December 2019 to 23 February 2020. About the comparison of mild and severe patients, 15 studies^[1, 10-23]^ described patient characteristics, 15 studies described comorbidities^[1, 10-17, 20-25]^, 8 studies^[1, 10, 14, 17, 19, 20, 22, 23]^ described vital signs, 17 studies compared symptoms^[1, 10-12, 14-26]^ and 19 studies^[1, 10-27]^ described laboratory findings. Guan, etc’study included 1099 patients from 552 hospitals in 30 provinces, and described the clinical characteristics of mild and severe patients in detail. Because of this article may be overlaps in population data with other studies, and the results are mainly described using median and quartile intervals, so we did not include the data in our meta-analysis, and mainly used descriptive analysis to compare the results with our studies.

**Table 1.**
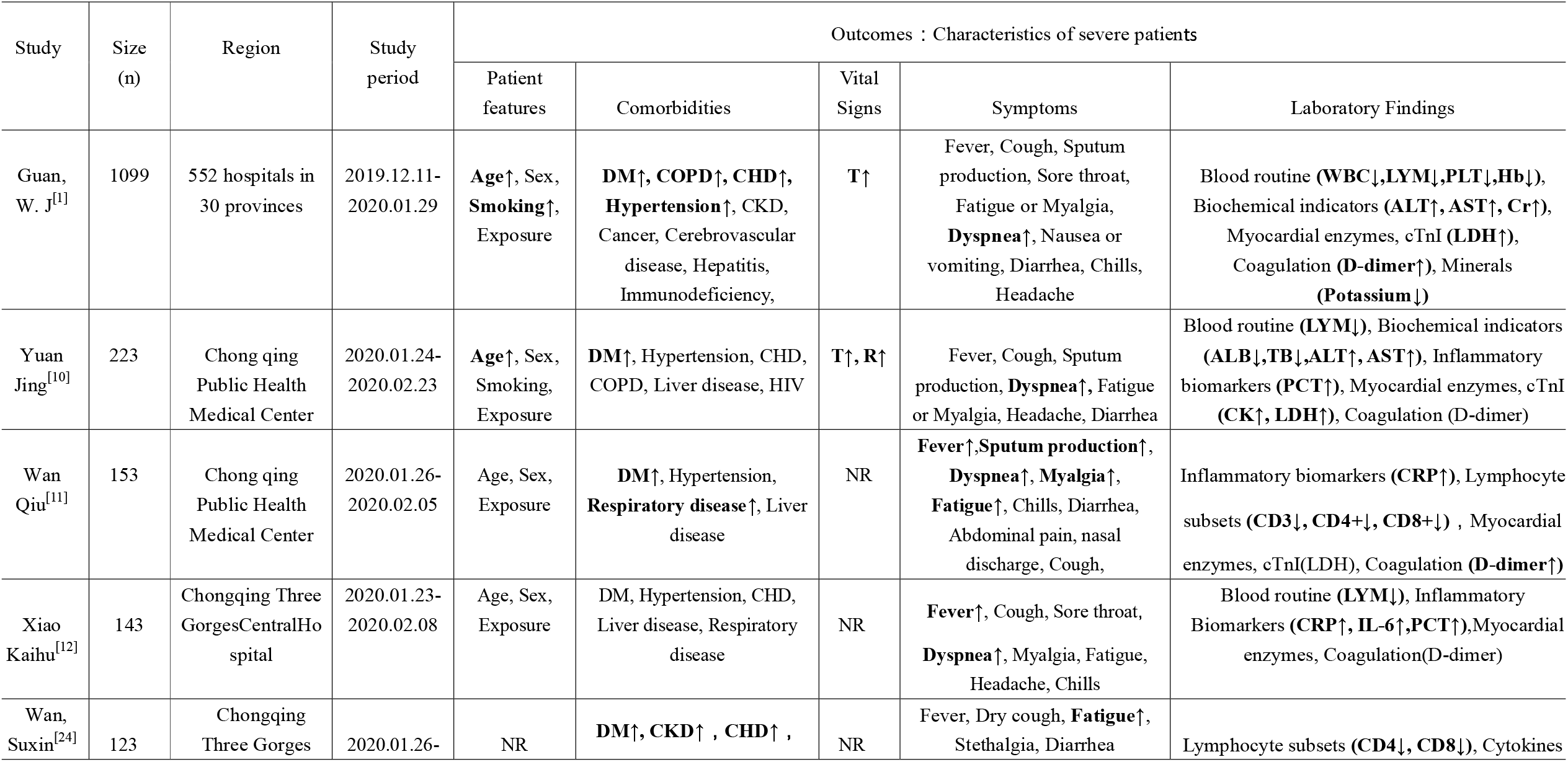
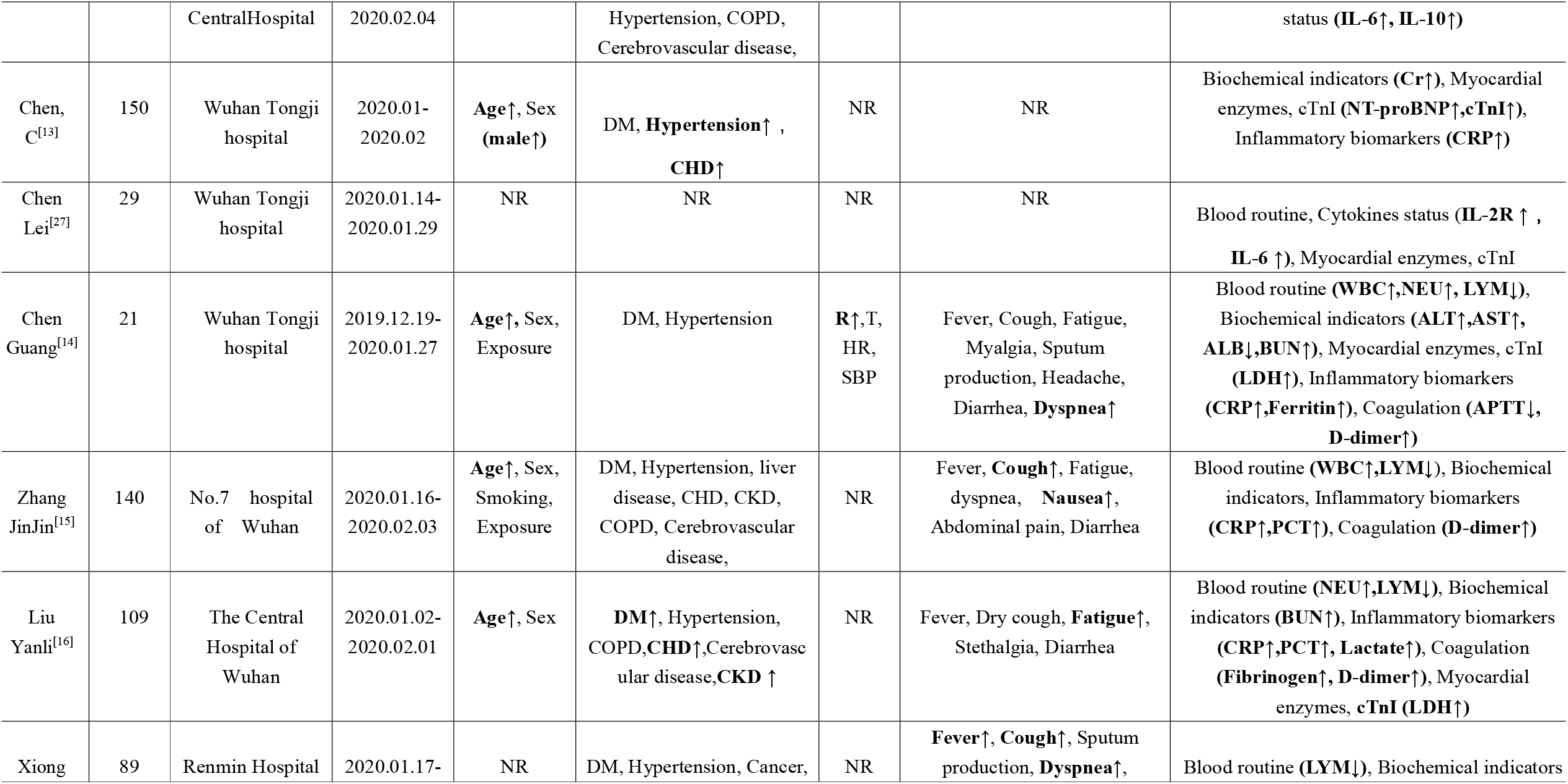
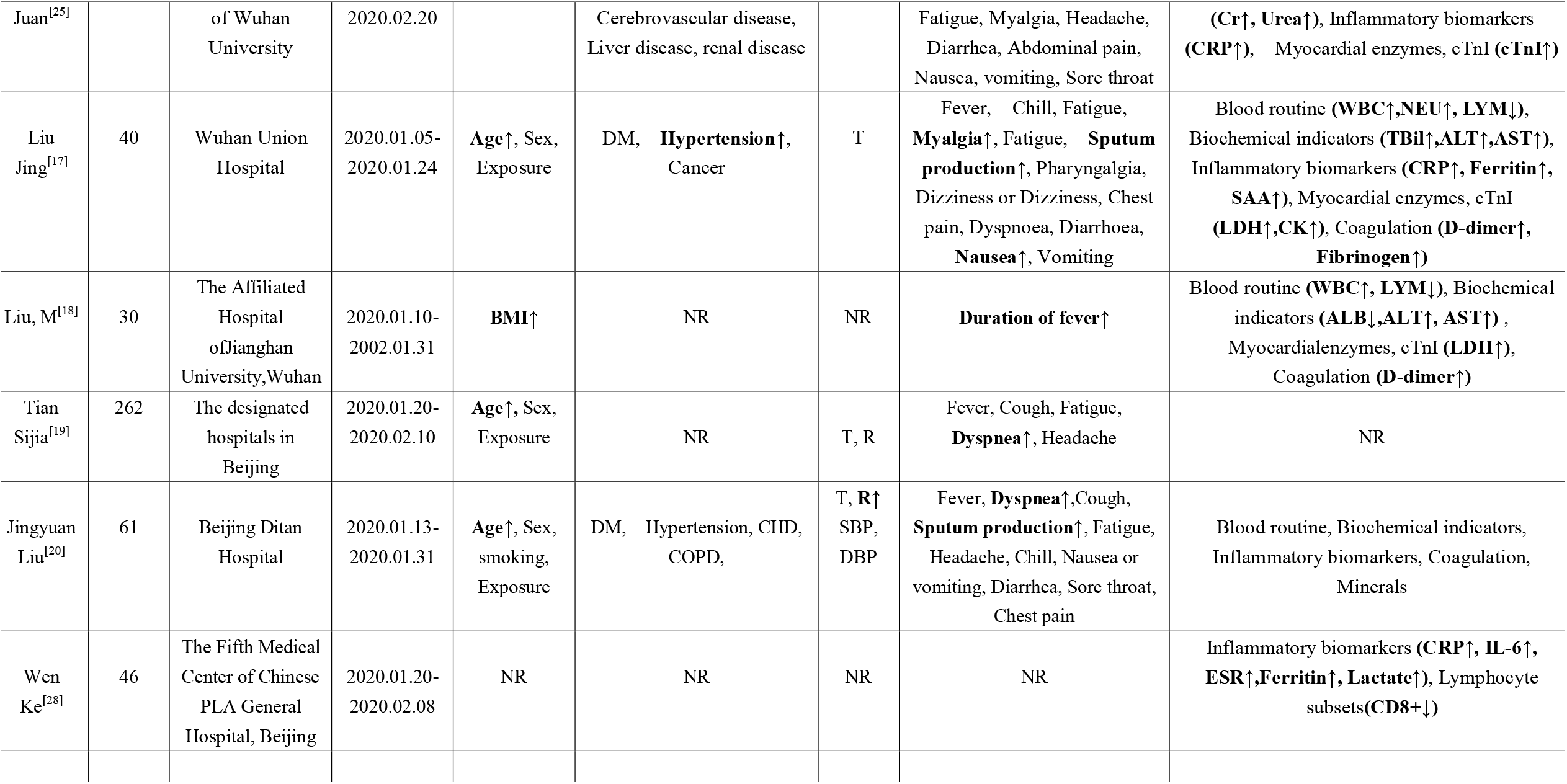
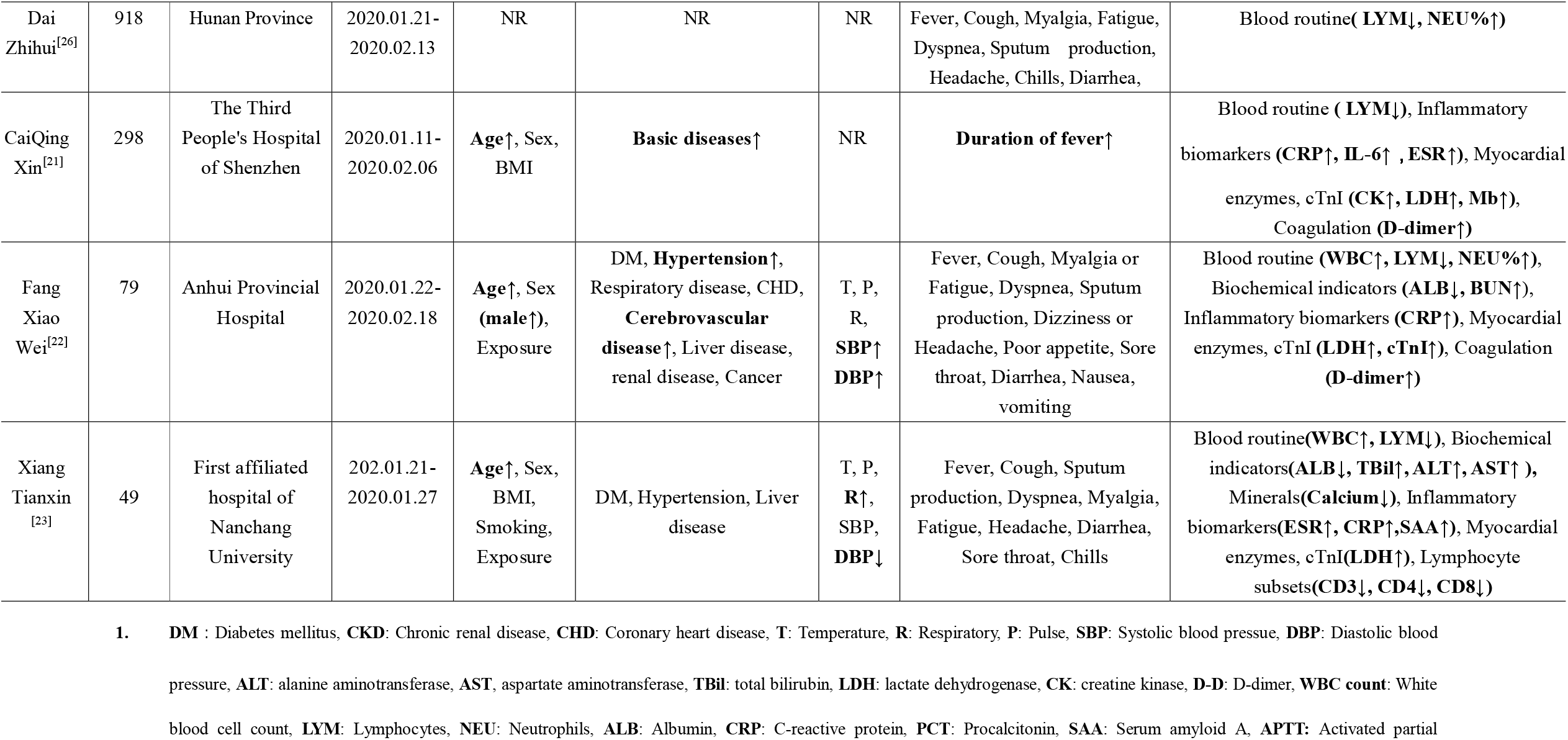
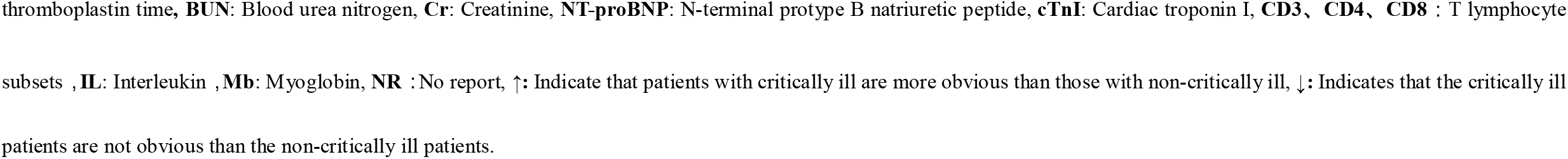
Summary the characteristics of 20 studies that described the risk factors with COVID-19 patients^1^.

| Study | Size (n) | Region | Study period | Outcomes : Characteristics of severe patients |  |  |  |  |
| --- | --- | --- | --- | --- | --- | --- | --- | --- |
|  |  |  |  | Patient features | Comorbidities | Vital Signs | Symptoms | Laboratory Findings |
| Guan, W. J<sup>[1]</sup> | 1099 | 552 hospitals in 30 provinces | 2019.12.11-2020.01.29 | **Age**↑, Sex, **Smoking**↑, Exposure | **DM**↑, **COPD**↑, **CHD**↑, **Hypertension**↑, CKD, Cancer, Cerebrovascular disease, Hepatitis, Immunodeficiency, | **T**↑ | Fever, Cough, Sputum production, Sore throat, Fatigue or Myalgia, **Dyspnea**↑, Nausea or vomiting, Diarrhea, Chills, Headache | Blood routine (**WBC**↓,**LYM**↓,**PLT**↓,**Hb**↓), Biochemical indicators (**ALT**↑, **AST**↑, **Cr**↑), Myocardial enzymes, cTnI (**LDH**↑), Coagulation (**D-dimer**↑), Minerals (**Potassium** ↓) |
| Yuan Jing<sup>[10]</sup> | 223 | Chong qing Public Health Medical Center | 2020.01.24-2020.02.23 | **Age**↑, Sex, Smoking, Exposure | **DM**↑, Hypertension, CHD, COPD, Liver disease, HIV | **T**↑, **R**↑ | Fever, Cough, Sputum production, **Dyspnea**↑, Fatigue or Myalgia, Headache, Diarrhea | Blood routine (**LYM**↓), Biochemical indicators (**ALB**↓,**TB**↓,**ALT**↑, **AST**↑), Inflammatory biomarkers (**PCT**↑), Myocardial enzymes, cTnI (**CK**↑, **LDH**↑), Coagulation (D-dimer) |
| Wan Qiu<sup>[11]</sup> | 153 | Chong qing Public Health Medical Center | 2020.01.26-2020.02.05 | Age, Sex, Exposure | **DM**↑, Hypertension, **Respiratory disease**↑, Liver disease | NR | **Fever**↑,**Sputum production**↑, **Dyspnea**↑, **Myalgia**↑, **Fatigue**↑, Chills, Diarrhea, Abdominal pain, nasal discharge, Cough, | Inflammatory biomarkers (**CRP**↑), Lymphocyte subsets (**CD3**↓, **CD4**+↓, **CD8**+↓) , Myocardial enzymes, cTnI(LDH), Coagulation (**D-dimer**↑) |
| Xiao Kaihu<sup>[12]</sup> | 143 | Chongqing Three GorgesCentralHospital | 2020.01.23-2020.02.08 | Age, Sex, Exposure | DM, Hypertension, CHD, Liver disease, Respiratory disease | NR | **Fever**↑, Cough, Sore throat, **Dyspnea**↑, Myalgia, Fatigue, Headache, Chills | Blood routine (**LYM**↓), Inflammatory Biomarkers (**CRP**↑, **IL-6**↑,**PCT**↑),Myocardial enzymes, Coagulation(D-dimer) |
| Wan, Suxin<sup>[24]</sup> | 123 | Chongqing Three Gorges | 2020.01.26- | NR | **DM**↑, **CKD**↑ , **CHD**↑ , | NR | Fever, Dry cough, **Fatigue**↑, Stethalgia, Diarrhea | Lymphocyte subsets (**CD4**↓, **CD8**↓), Cytokines |
|  |  | CentralHospital | 2020.02.04 |  | Hypertension, COPD, Cerebrovascular disease, |  |  | status (**IL-6**↑, **IL-10**↑) |
| Chen, C<sup>[13]</sup> | 150 | Wuhan Tongji hospital | 2020.01-2020.02 | **Age**↑, Sex (**male**↑) | DM, **Hypertension**↑ ,<br>**CHD**↑ | NR | NR | Biochemical indicators (**Cr**↑), Myocardial enzymes, cTnI (**NT-proBNP**↑,**cTnI**↑), Inflammatory biomarkers (**CRP**↑) |
| Chen Lei<sup>[27]</sup> | 29 | Wuhan Tongji hospital | 2020.01.14-2020.01.29 | NR | NR | NR | NR | Blood routine, Cytokines status (**IL-2R** ↑ ,<br>**IL-6** ↑), Myocardial enzymes, cTnI |
| Chen Guang<sup>[14]</sup> | 21 | Wuhan Tongji hospital | 2019.12.19-2020.01.27 | **Age**↑, Sex, Exposure | DM, Hypertension | **R**↑,T, HR, SBP | Fever, Cough, Fatigue, Myalgia, Sputum production, Headache, Diarrhea, **Dyspnea**↑ | Blood routine (**WBC**↑,**NEU**↑, **LYM**↓), Biochemical indicators (**ALT**↑,**AST**↑, **ALB**↓,**BUN**↑), Myocardial enzymes, cTnI (**LDH**↑), Inflammatory biomarkers (**CRP**↑,**Ferritin**↑), Coagulation (**APTT**↓, **D-dimer**↑) |
| Zhang JinJin<sup>[15]</sup> | 140 | No.7 hospital of Wuhan | 2020.01.16-2020.02.03 | **Age**↑, Sex, Smoking, Exposure | DM, Hypertension, liver disease, CHD, CKD, COPD, Cerebrovascular disease, | NR | Fever, **Cough**↑, Fatigue, dyspnea, **Nausea**↑, Abdominal pain, Diarrhea | Blood routine (**WBC**↑,**LYM**↓), Biochemical indicators, Inflammatory biomarkers (**CRP**↑,**PCT**↑), Coagulation (**D-dimer**↑) |
| Liu Yanli<sup>[16]</sup> | 109 | The Central Hospital of Wuhan | 2020.01.02-2020.02.01 | **Age**↑, Sex | **DM**↑, Hypertension, COPD,**CHD**↑,Cerebrovascular disease,**CKD** ↑ | NR | Fever, Dry cough, **Fatigue**↑, Stethalgia, Diarrhea | Blood routine (**NEU**↑,**LYM**↓), Biochemical indicators (**BUN**↑), Inflammatory biomarkers (**CRP**↑,**PCT**↑, **Lactate**↑), Coagulation (**Fibrinogen**↑, **D-dimer**↑), Myocardial enzymes, **cTnI** (**LDH**↑) |
| Xiong | 89 | Renmin Hospital | 2020.01.17- | NR | DM, Hypertension, Cancer, | NR | **Fever**↑, **Cough**↑, Sputum production, **Dyspnea**↑, | Blood routine (**LYM**↓), Biochemical indicators |
| Juan<sup>[25]</sup> |  | of Wuhan University | 2020.02.20 |  | Cerebrovascular disease, Liver disease, renal disease |  | Fatigue, Myalgia, Headache, Diarrhea, Abdominal pain, Nausea, vomiting, Sore throat | (**Cr**↑, **Urea**↑), Inflammatory biomarkers (**CRP**↑), Myocardial enzymes, cTnI (**cTnI**↑) |
| Liu Jing<sup>[17]</sup> | 40 | Wuhan Union Hospital | 2020.01.05-2020.01.24 | **Age**↑, Sex, Exposure | DM, **Hypertension**↑, Cancer | T | Fever, Chill, Fatigue, **Myalgia**↑, Fatigue, **Sputum production**↑, Pharyngalgia, Dizziness or Dizziness, Chest pain, Dyspnoea, Diarrhoea, **Nausea**↑, Vomiting | Blood routine (**WBC**↑, **NEU**↑, **LYM**↓), Biochemical indicators (**TBil**↑, **ALT**↑, **AST**↑), Inflammatory biomarkers (**CRP**↑, **Ferritin**↑, **SAA**↑), Myocardial enzymes, cTnI (**LDH**↑, **CK**↑), Coagulation (**D-dimer**↑, **Fibrinogen**↑) |
| Liu, M<sup>[18]</sup> | 30 | The Affiliated Hospital of Jiangnan University, Wuhan | 2020.01.10-2020.01.31 | **BMI**↑ | NR | NR | **Duration of fever**↑ | Blood routine (**WBC**↑, **LYM**↓), Biochemical indicators (**ALB**↓, **ALT**↑, **AST**↑), Myocardial enzymes, cTnI (**LDH**↑), Coagulation (**D-dimer**↑) |
| Tian Sijia<sup>[19]</sup> | 262 | The designated hospitals in Beijing | 2020.01.20-2020.02.10 | **Age**↑, Sex, Exposure | NR | T, R | Fever, Cough, Fatigue, **Dyspnea**↑, Headache | NR |
| Jingyuan Liu<sup>[20]</sup> | 61 | Beijing Ditan Hospital | 2020.01.13-2020.01.31 | **Age**↑, Sex, smoking, Exposure | DM, Hypertension, CHD, COPD, | T, **R**↑ SBP, DBP | Fever, **Dyspnea**↑, Cough, **Sputum production**↑, Fatigue, Headache, Chill, Nausea or vomiting, Diarrhea, Sore throat, Chest pain | Blood routine, Biochemical indicators, Inflammatory biomarkers, Coagulation, Minerals |
| Wen Ke<sup>[28]</sup> | 46 | The Fifth Medical Center of Chinese PLA General Hospital, Beijing | 2020.01.20-2020.02.08 | NR | NR | NR | NR | Inflammatory biomarkers (**CRP**↑, **IL-6**↑, **ESR**↑, **Ferritin**↑, **Lactate**↑), Lymphocyte subsets (**CD8+**↓) |
| Dai Zhihui<sup>[26]</sup> | 918 | Hunan Province | 2020.01.21-2020.02.13 | NR | NR | NR | Fever, Cough, Myalgia, Fatigue, Dyspnea, Sputum production, Headache, Chills, Diarrhea, | Blood routine( **LYM**↓, **NEU**%↑) |
| CaiQing Xin<sup>[21]</sup> | 298 | The Third People's Hospital of Shenzhen | 2020.01.11-2020.02.06 | **Age**↑, Sex, BMI | **Basic diseases**↑ | NR | **Duration of fever**↑ | Blood routine ( **LYM**↓), Inflammatory biomarkers (**CRP**↑, **IL-6**↑ , **ESR**↑), Myocardial enzymes, cTnI (**CK**↑, **LDH**↑, **Mb**↑), Coagulation (**D-dimer**↑) |
| Fang Xiao Wei<sup>[22]</sup> | 79 | Anhui Provincial Hospital | 2020.01.22-2020.02.18 | **Age**↑, Sex (**male**↑), Exposure | DM, **Hypertension**↑, Respiratory disease, CHD, **Cerebrovascular disease**↑, Liver disease, renal disease, Cancer | T, P, R, **SBP**↑ **DBP**↑ | Fever, Cough, Myalgia or Fatigue, Dyspnea, Sputum production, Dizziness or Headache, Poor appetite, Sore throat, Diarrhea, Nausea, vomiting | Blood routine (**WBC**↑, **LYM**↓, **NEU**%↑), Biochemical indicators (**ALB**↓, **BUN**↑), Inflammatory biomarkers (**CRP**↑), Myocardial enzymes, cTnI (**LDH**↑, **cTnI**↑), Coagulation (**D-dimer**↑) |
| Xiang Tianxin<sup>[23]</sup> | 49 | First affiliated hospital of Nanchang University | 202.01.21-2020.01.27 | **Age**↑, Sex, BMI, Smoking, Exposure | DM, Hypertension, Liver disease | T, P, **R**↑, SBP, **DBP**↓ | Fever, Cough, Sputum production, Dyspnea, Myalgia, Fatigue, Headache, Diarrhea, Sore throat, Chills | Blood routine(**WBC**↑, **LYM**↓), Biochemical indicators(**ALB**↓, **TBil**↑, **ALT**↑, **AST**↑ ), Minerals(**Calcium**↓), Inflammatory biomarkers(**ESR**↑, **CRP**↑, **SAA**↑), Myocardial enzymes, cTnI(**LDH**↑), Lymphocyte subsets(**CD3**↓, **CD4**↓, **CD8**↓) |

1. **DM** : Diabetes mellitus, **CKD**: Chronic renal disease, **CHD**: Coronary heart disease, **T**: Temperature, **R**: Respiratory, **P**: Pulse, **SBP**: Systolic blood pressue, **DBP**: Diastolic blood pressure, **ALT**: alanine aminotransferase, **AST**, aspartate aminotransferase, **TBil**: total bilirubin, **LDH**: lactate dehydrogenase, **CK**: creatine kinase, **D-D**: D-dimer, **WBC count**: White blood cell count, **LYM**: Lymphocytes, **NEU**: Neutrophils, **ALB**: Albumin, **CRP**: C-reactive protein, **PCT**: Procalcitonin, **SAA**: Serum amyloid A, **APTT**: Activated partial
thromboplastin time, **BUN**: Blood urea nitrogen, **Cr**: Creatinine, **NT-proBNP**: N-terminal protype B natriuretic peptide, **cTnI**: Cardiac troponin I, **CD3**, **CD4**, **CD8** : T lymphocyte subsets , **IL**: Interleukin , **Mb**: Myoglobin, **NR** : No report, ↑: Indicate that patients with critically ill are more obvious than those with non-critically ill, ↓: Indicates that the critically ill patients are not obvious than the non-critically ill patients.

### Quality assessment

The quality of included studies was assessed by the observational study quality evaluation criteria recommended by AHRQ. All studies included clear data sources, inclusion and exclusion criteria, and reasonable control of confounding factors. However, only a few studies clearly reported the quality control of the study and the treatment of missing data (See **Appendix table 1**).

### Meta-analysis

The results of the meta-analysis are shown in **Table 2**. More intuitive results can be found in the forest-plot **(Figure 2, Figure 3)**.

**Table 2.** The meta-analysis of risk factors for severe patients with COVID-19.

| **Risk factor** | **Definition** | **Number of studies** | **Size (n)** | **OR/WMD/SMD (CI 95%)** | **I<sup>2</sup>** |
| --- | --- | --- | --- | --- | --- |
| **Patient characteristics** |  |  |  |  |  |
| Age (year) | Continuous | 6 | 681 | 11.89 [8.63, 15.14] | 54.40% |
| Gender | Male vs. Female | 10 | 1494 | 1.61 [1.25, 2.07] | 0% |
| BMI (kg/m<sup>2</sup>) | Continuous | 2 | 79 | 3.38 [0.07, 6.69] | 67.20% |
| Smoking | Yes vs. No | 3 | 412 | 1.4 [0.65, 3.01] | 0% |
| **Comorbidities** |  |  |  |  |  |
| Diabetes | Yes vs. No | 10 | 1083 | 3.04 [2.01, 4.60] | 20.40% |
| Hypertension | Yes vs. No | 10 | 1083 | 2.31 [1.68, 3.18] | 47.10% |
| Cardiovascular disease | Yes vs. No | 7 | 906 | 2.76 [1.39, 5.45] | 25.70% |
| Chronic obstructive pulmonary disease | Yes vs. No | 5 | 623 | 3.56 [1.33, 9.54] | 0% |
| **Vital Signs** |  |  |  |  |  |
| Respiratory rate (per min ) | Continuous | 2 | 70 | 5.29 [2.56, 8.01] | 42.80% |
| **Symptoms** |  |  |  |  |  |
| Fever | Yes vs. No | 10 | 2032 | 2.11 [1.11, 4.02] | 64.20% |
| Dyspnea | Yes vs. No | 8 | 977 | 8.83 [2.82, 27.67] | 79.10% |
| **Laboratory Findings** |  |  |  |  |  |
| **Blood routine** |  |  |  |  |  |
| White blood cell count(×10<sup>9</sup>/L) | Continuous | 7 | 595 | 1.76 [0.31, 3.22] | 88.70% |
| Lymphocytes count (×10<sup>9</sup>/L) | Continuous | 6 | 563 | -0.42 [-0.52, -0.33] | 11.50% |
| Neutrophils count (×10<sup>9</sup>/L) | Continuous | 2 | 159 | 2.92 [-1.33, 7.17] | 93% |
| **Biochemical indicators** |  |  |  |  |  |
| Albumin(g/L) | Continuous | 4 | 323 | -5.74 [-7.94, -3.54] | 62.80% |
| Alanine aminotransferase(IU/L) | Continuous | 4 | 189 | 16.97 [2.18, 31.76] | 89.10% |
| Aspartate aminotransferase(IU/L) | Continuous | 4 | 229 | 20.60 [6.81, 34.40] | 86.90% |
| Lactate dehydrogenase(umol/L) | Continuous | 3 | 120 | 3.93 [2.01, 5.86] | 0% |
| Creatinine (umol/L) | Continuous | 5 | 422 | 8.32 [-1.63, 18.28] | 62.10% |
| **Inflammatory biomarkers** |  |  |  |  |  |
| C-reactive protein(SMD) | Continuous | 5 | 463 | 2.11 [0.71, 3.51] | 96.10% |
| Procalcitonin(ng/ml) | Continuous | 5 | 455 | 0.16 [0.04, 0.28] | 77.80% |
| **Myocardial enzymes** |  |  |  |  |  |
| Lactate dehydrogenase(SMD) | Continuous | 5 | 291 | 2.0 [1.20, 2.80] | 81.10% |
| Creatine kinase(IU/L) | Continuous | 3 | 213 | 23.55 [17.08, 30.02] | 30.80% |
| **Coagulation** |  |  |  |  |  |
| D-dimer (mg/L) | Continuous | 5 | 396 | 0.67 [0.02, 1.32] | 72% |

**Figure 2.**
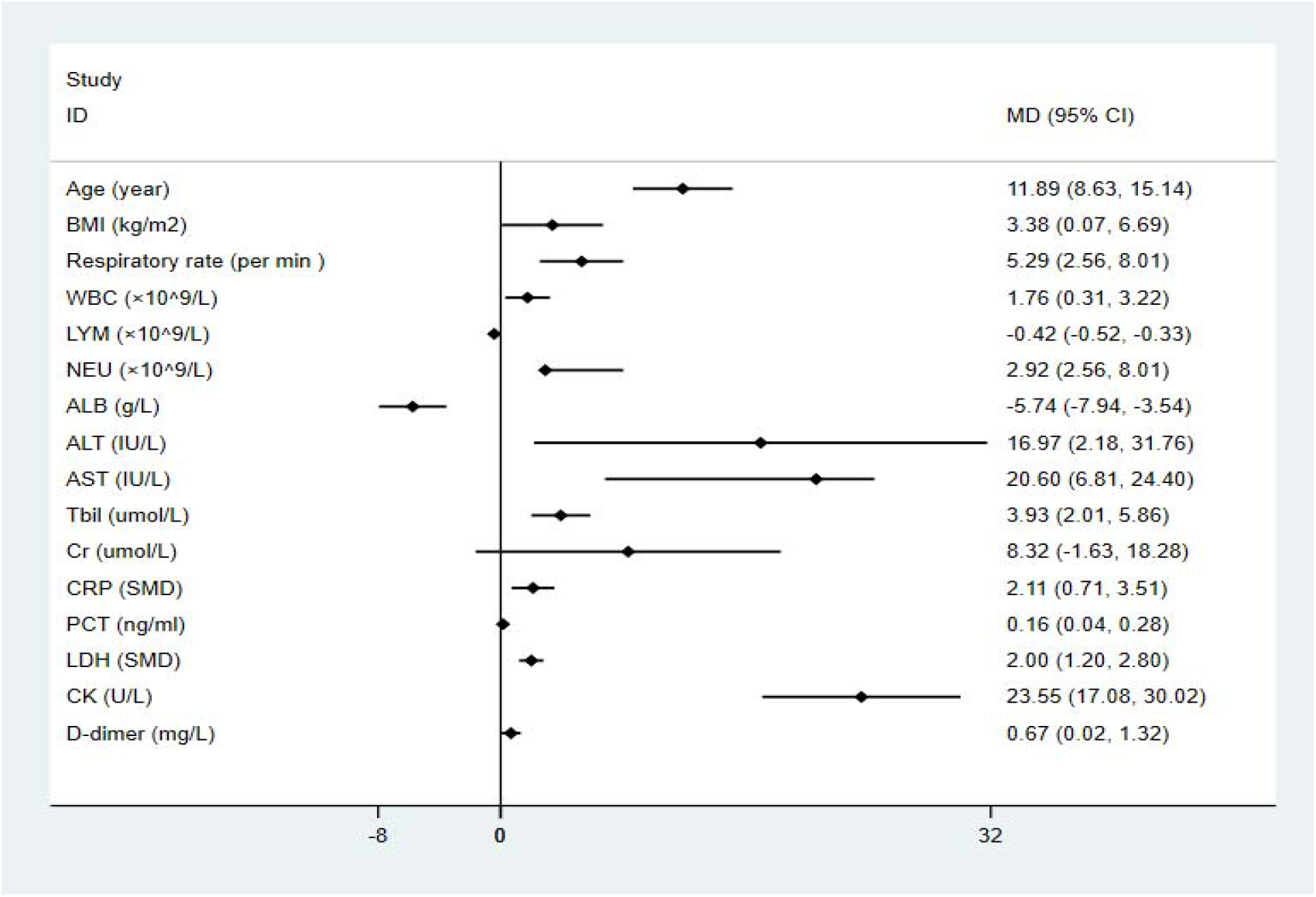
The forest plot of risk factors with COVID-19 patients on continuous variable.

**Figure 3.**
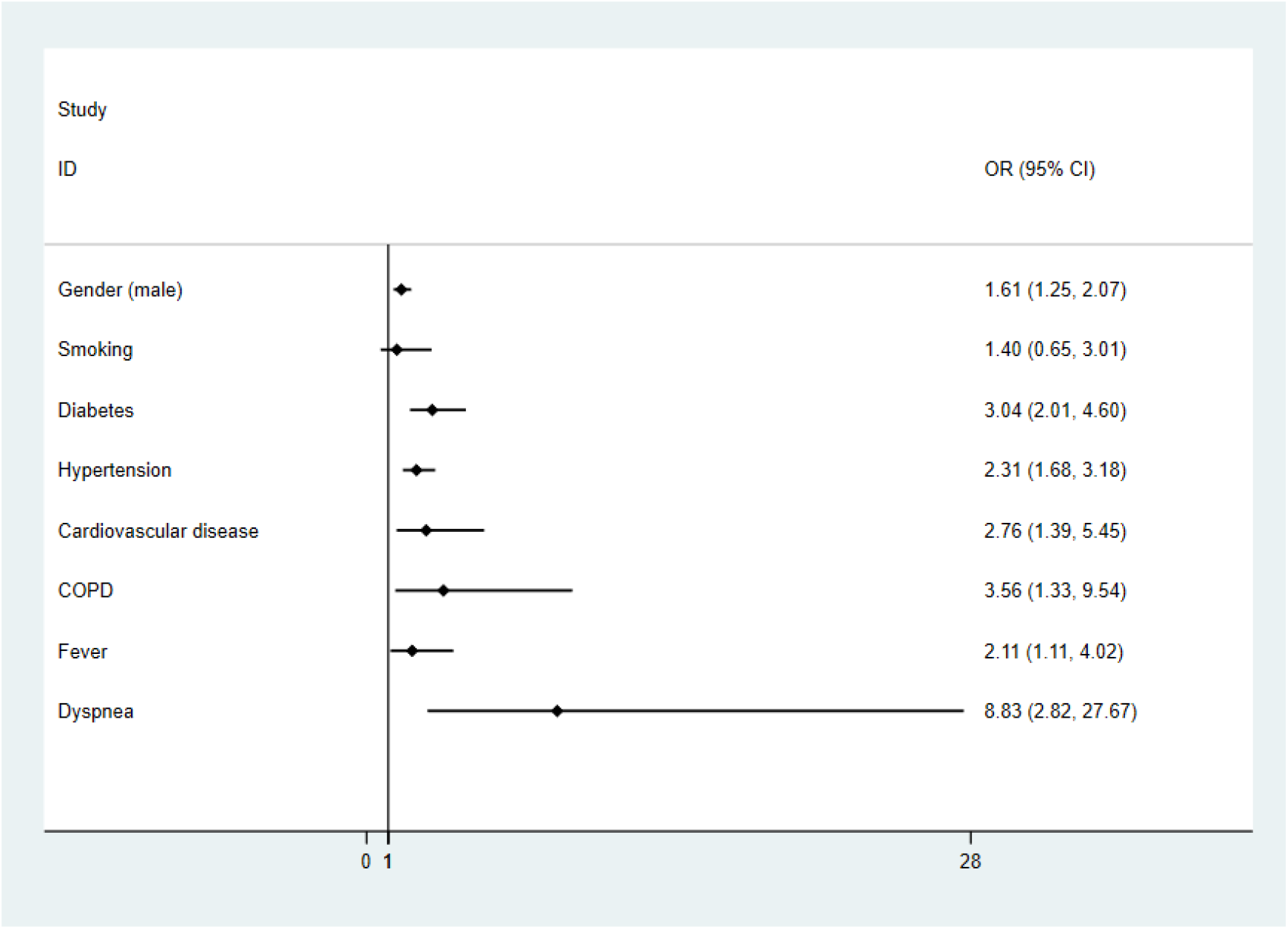
The forest plot of risk factors with COVID-19 patients onbinary variable.

**Figure 4.**
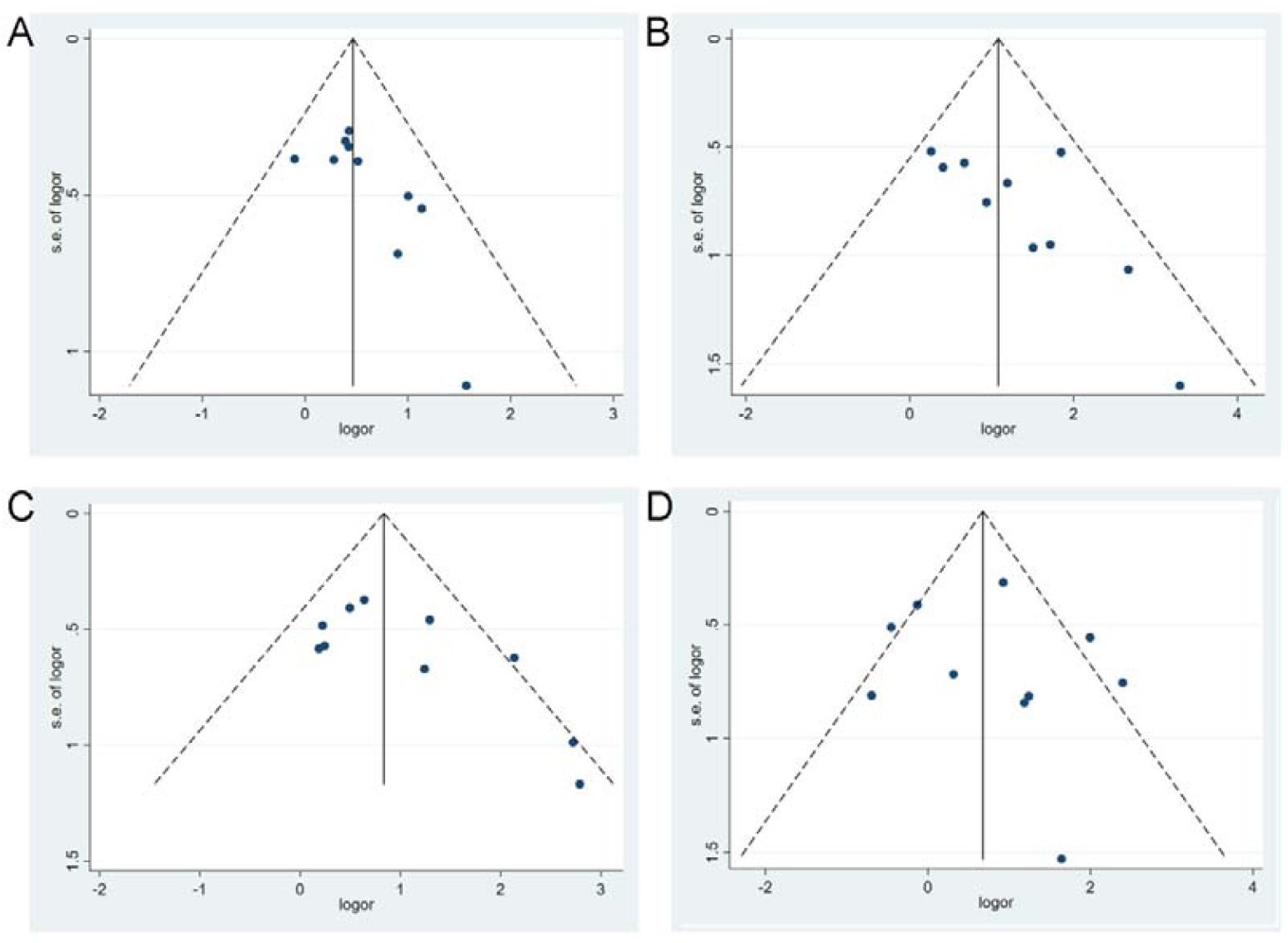
The funnel-plots of A (Gender), B (Diabetes), C (Hypertension), D (Fever).

### Patient characteristics (Age, Gender, BMI and Smoking)

A random-effects model (***I***^***2***^=54.4%) was used on 6 articles^[10, 12, 13, 17, 22, 23]^ involving 681 patients to analyze the risk factors of age for severe patients with COVID-19. Compared with non-severe patients, the average age of severe patients was older (WMD=11.89 [8.63, 15.14]) than mild patients. Results about gender on ten studies^[10, 12, 13, 15-17, 19, 21-23]^ included 1494 patients suggesting that men were more likely to have severe pneumonia than women (OR=1.61 [1.25, 2.07]). Only have two studies^[18, 23]^ on BMI included 79 patients showed that the BMI value of severe patients was higher than non-severe patients (WMD = 3.38 [0.07, 6.69]). A fixed-effects model was used on 3 articles^[10, 15, 23]^ involving 412 patients to analyze the risk factors of smoking showed that there was no significant correlation between smoking and the severe pneumonia (OR = 1.4 [0.65, 3.01]).

### Comorbidities (Diabetes, Hypertension, Coronary heart disease, Chronic obstructive pulmonary disease)

Our research show that patients with the following underlying diseases are more likely to develop severe COVID-19: Diabetes mellitus (DM):OR = 3.04 [2.01, 4.60], Hypertension (HTN):OR = 2.31 [1.68, 3.18], Coronary heart disease (CHD):OR = 2.76 [1.39, 5.45], Chronic obstructive pulmonary disease (COPD):OR = 3.56 [1.33, 9.54]. All results were low heterogeneous **(*I***_***2***_<50%).

### Vital Signs (respiratory rate) and Symptoms (Fever and Dyspnea)

Regarding breathing, our study in 2 article^[14, 23]^ (included 70 patients) showing that patients with severe COVID-19 breathed faster (WMD = 5.29 [2.56, 8.01], I2 = 42.8%). Our study on fever (11 articles^[10, 12, 14-17, 19, 22, 23, 25, 26]^) and dyspnea (8 articles^[10, 12, 14, 15, 17, 19, 22, 25]^) founded that fever and dyspnea may be the risk factors for severe patients (Fever: OR = 2.11 [1.11, 4.02]; Dyspnea: OR = 8.83 [2.82,27.67]).

### Laboratory Findings (Blood routine, Biochemical indicators, Inflammatory biomarkers and Coagulation)

Our study in laboratory findings showed that white-cell count (WMD=1.76 [0.31, 3.22]) increased in severe patients, while the proportion of lymphocytes decreased (WMD=-0.42 [-0.52, -0.33]). Compared with non-severe patients, severe patients have higher alanine aminotransferase (ALT), aspartate aminotransferase (AST), and total bilirubin (Tbil) (ALT: WMD = 16.97 [2.18, 31.76]), AST: WMD = 20.60 [6.81, 34.40], Tbil: WMD = 3.93 [2.01, 5.86]), but creatinine (Cr), albumin (ALB) and neutrophil count (N count) does not seem obvious difference (Cr: WMD = 8.32 [-1.63, 18.28], ALB:WMD = -5.74 [-7.94, -3.54], N count, WMD=2.92 [-1.33, 7.17]). Regarding of C-reactive protein (CRP) and Procalcitonin (PCT), severe patients were higher than non-severe patients (CRP: SMD = 2.11 [0.71, 3.51], PCT: WMD = 0.16 [0.04, 0.28]). What’more, our study found that compared with non-severe patients, lactate dehydrogenase (LDH), creatine kinase (CK) and D-dimer (D-D) increased more significantly in severe patients (LDH: SMD = 2.0 [1.20, 2.80], CK: WMD = 23.55 [17.08, 30.02], D-D: WMD = 0.67 [0.02, 1.32]).

### Publication bias

We only draw a funnel-plots for the outcome indicators with more than 10 studies, and judge the publication bias of the results by observing the symmetry of the funnel-plots. As shown, our funnel-plots is roughly symmetrical, indicating that publication bias is small. In addition, our research not only includes published studies, but for unpublished literature on medRxiv and bioxiv, as long as it meets our inclusion criteria, we also include them, aim to minimize publication bias.

## Discussion

This study shows the conclusive evidence as risk factors for severe COVID-19 patients, such as **patient characteristics (**age, gender, BMI**), comorbidities (**DM, HTN, CHD, COPD**), vital signs (**respiratory rate**), symptoms (**fever and dyspnea) and **laboratory findings (**blood routine, biochemical indicators, inflammatory biomarkers and coagulation).

At present, no special drug for SARS-CoV-2 infection has been found and the research process of the vaccine is still in clinical trials, the treatment of COVID-19 is mainly by Symptomatic treatment. Based on Chinese experience, mild patients and their close contacts treated in close isolation and follow-up are likely to be sufficient to manage the disease^[29-31]^. But for severe patients, intensive care and aggressive treatments are needed. By searching various databases, we found that this is the first article of systematic review and meta-analysis to summarize the characteristics of severely infected patients with SARS-CoV-2.

The article^[1]^ of Guan, etc shows that the Median (IQR) age of critically ill patients was 52.0 (40.0–65.0) which was older than the non-critically ill patients whose median (IQR) age was 45.0 (34.0–57.0). Similarly in our review, we found that patients with critically ill were older than those with non-critically ill patients, and most patients are male. Moreover, the presence of coexisting illness (such as DM, HTN, CHD, COPD) was more common among patients with severe disease than among those with nonsevere disease. We all know that the elderly with basic diseases, especially those with diabetes usually have a high blood glucose status for a long time, so their ability to defend against infection is low^[32]^. They are at high risk for various infections, and once infected, they can easily develop into severe diseases. The lower sensitivity of women to viral infections may be due to the protection from X chromosomes and sex hormones, which play an important role in innate and adaptive immunity^[33]^. So, we think that elderly male patients with underlying diseases are at high risk for severely ill with COVID-19 and needs special attention from clinicians. In addition, our study also found that patients with high BMI are more likely to develop into severe pneumonia, which may be related to the high expression of angiotensin-converting enzyme 2 (ACE2) in obese patients^[34]^. We know that ACE2 is a binding receptor for SARS-CoV-2 and has a very high affinity between them. Obese patients are richer in adipose tissue, have a larger total amount of ACE2 receptors^[34]^, and are more susceptible to SARS-CoV-2. It is suggested that obese people are at high risk of COVID-19, and they are the key protection targets in epidemic prevention work.

The symptoms of COVID-19 include fever, cough, sputum production, sore throat, fatigue or myalgia, dyspnea, nausea or vomiting, diarrhea, chills and headache, etc. In most studies, the symptom of fever and dyspnea occur more frequently in critically ill patients^[1, 11, 12]^, which is consistent with our study. regarding of vital signs, we found that critically ill patients usually breathed more faster than non-critically ill patients, which related to the low level of blood oxygen saturation caused by severe lung disease in critically ill patients. A study^[23]^ from 49 people found no significant difference in pulse rate between severe patients and non-severe patients (90.6 ± 10.3 vs. 93.8 ± 13.7, p = 0.440), which is consistent with the findings of fang, et al^[22]^. At the same time, the fang’s study^[22]^ also found that the blood pressure of severe patients seems to be higher than that of non-severe patients [SBP: 133.3 ± 16.5 vs. 121.2 ± 9.5, p <0.001; DBP: 83.3 ± 11.7 vs. 74.2 ± 9.5, p <0.001]. while the study by Xiang, et al^[23]^ found no significant difference in systolic blood pressure (p = 0.769), but lower diastolic blood pressure (69.1 ± 10.8 vs. 80.7 ± 12.7, p = 0.013) compared with non-severe patients.

As for laboratory tests, patients with severe disease had more prominent laboratory abnormalities (including lymphocytopenia, hypoalbuminemia, elevated levels of ALT, AST, Tbil, LDH, CK, etc) than those with nonsevere disease. Many previous studies have reported lymphocytopenia in patients with COVID-19^[35]^, lymphocytopenia is a significant feature of severe patients with SARS-CoV-2 infection because targeted invasion by SARS-CoV viral particles can damages thecytoplasmic component of the lymphocyte and cause its necrosis or apoptosis^[36]^. In this meta-analysis, we found that SARS-CoV-2 can damage the various organs throughout the body in addition to the lung, manifested as the elevated levels of ALT, AST, Tbil, LDH, CK, etc in different degree. As we know, coronavirus (CoV) is a pathogen that infects the respiratory, gastrointestinal, liver, and central nervous systems of humans and many other wild animals^[37, 38]^. SARS-CoV-2, also known as the sister virus of SARS-CoV, has 85% nucleotide homology^[39, 40]^. The latest pathology reports^[41]^ found that the pathological manifestations of SARS-CoV-2 and SARS-CoV are similar, mainly manifesting as acute respiratory distress syndrome (ARDS) and multiple organ failure, which also explains the cause of multiple organ dysfunction in critically ill patients. SARS-CoV-2 has been shown to infect human respiratory epithelial cells through the angiotensin-converting enzyme 2 receptors on human cells^[42, 43]^. Many studies^[34, 44]^ have found that the levels of ACE2 RNA was higher in heart, kidney, intestinal tract, gallbladder, adipose tissue and testicles than lungs in human body, which also explains the reason why multi-organ dysfunction is prone to occur in severe patients.

Inflammatory biomarker is also a common feature in the patients with COVID-19 and might be a critical factor associated with disease severity and mortality. In the study, we found that CRP and PCT increased in varying degrees for severe patients. Some research also reported the elevated level of ESR, IL-6, and IL-10 in critically ill patients^[24]^. It is considered that multiple cytokines are secreted after the infection of microorganisms causing a strong inflammatory response and the damage of immune system, indicated that critically ill patients may have more severe systemic inflammatory response, so attention should be paid to strengthen the treatment of anti-inflammatory^[23]^.

## Limitations

However, there are still some limitations lying in our study. First, all included studies were cross-sectional, precluding the possibility to establish inferences regarding causality. Second, populations, risk factors analyzed, lengths of follow-up, and statistical methods might differ and cause study heterogeneity. Third, although this study reviewed the risk factors of severe COVID-19 patients and their impact, assessing the effect size of each risk factor and developing risk models were important.

## Conclusion

The severity of SARS-CoV-2 pneumonia has placed great pressure on intensive care resources in hospital, especially in some developing countries where lacking of medical staff and health resources, so early identification of severe patients and taking active and effective measures can reduce deaths.

## Data Availability

all data included in this study are available upon request by contact with the corresponding author.

## Acknowledgments

We thank all patients and their families involved in the study. Authors are also thankful to the Department of Endocrinology, Shengli Clinical Medical College of Fujian Medical University.

## Article Information

### Author Contributions

Gang Chen had full access to all of the data in the study and takes responsibility for the integrity of the data and the accuracy of the data analysis. Lizhen Xu and Yaqian Mao contributed to the study equally.

**Concept and design:** Lizhen Xu, Yaqian Mao, Gang Chen.

**Acquisition, analysis, or interpretation of data:** Lizhen Xu, Yaqian Mao

**Drafting of the manuscript:** Lizhen Xu, Yaqian Mao

**Critical revision of the manuscript for important intellectual content:** Lizhen Xu, Yaqian Mao, Gang Chen

**Statistical analysis:** Lizhen Xu

**Administrative, technical, or material support:** None

**Supervision:** Gang Chen

## Appendix

**Table 1.**
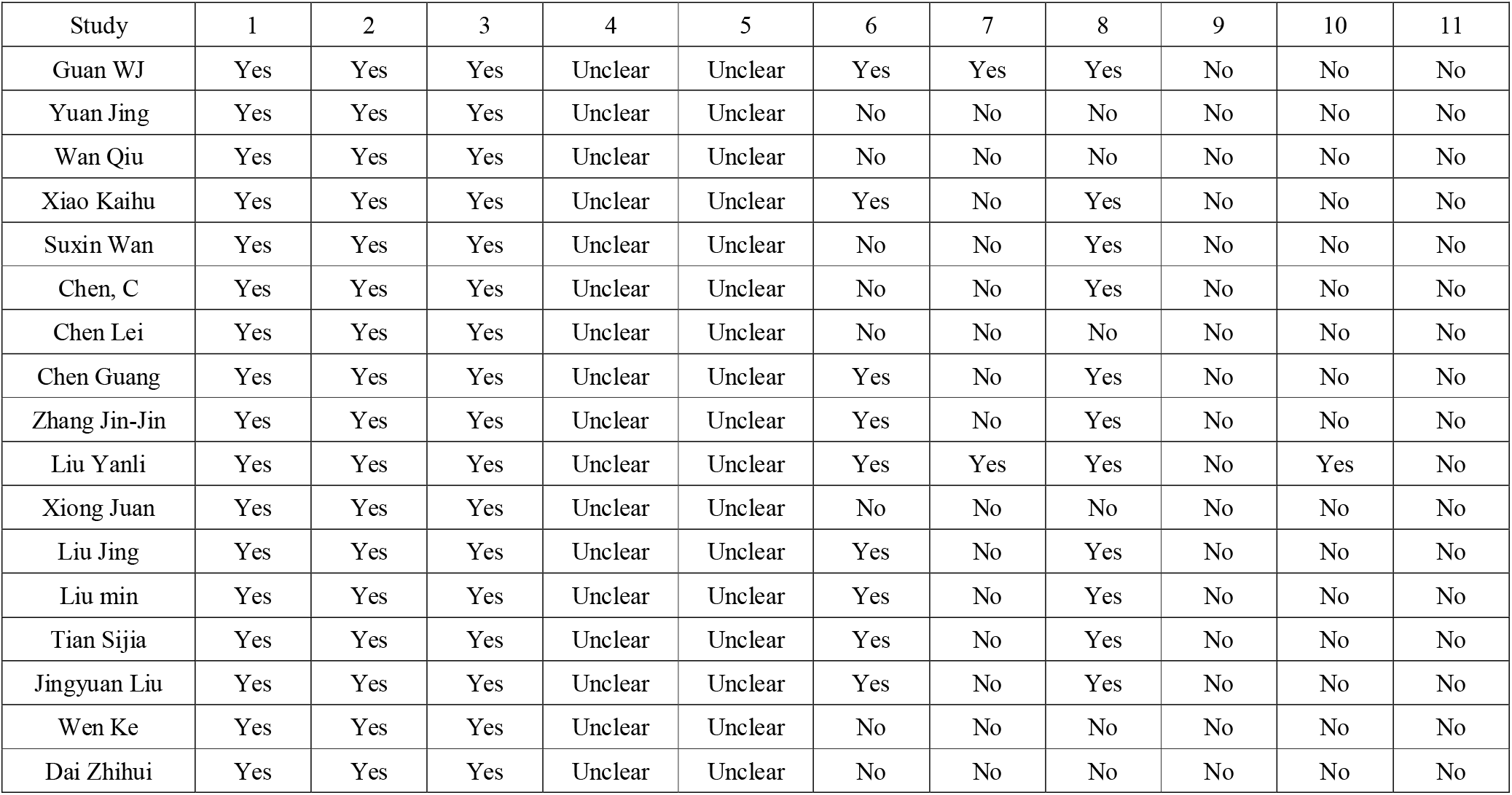
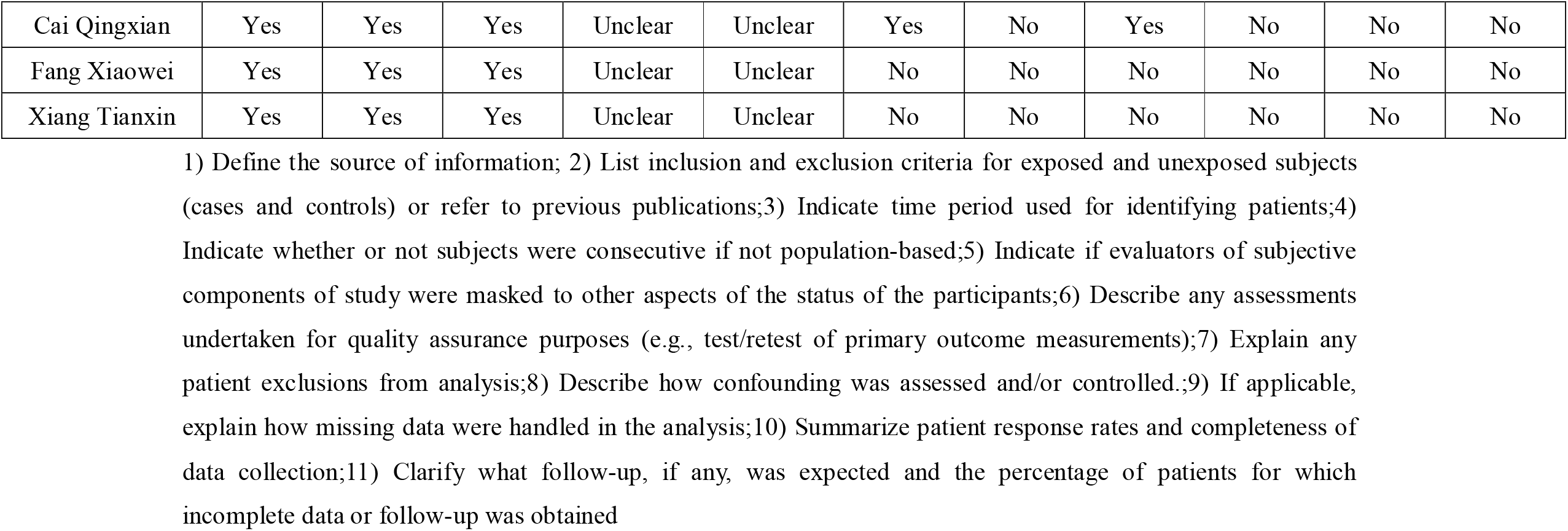
Quality assessment: Cross-Sectional/Prevalence Study Quality according to AHRQ.

## References

[1] Guan W J, Ni Z Y, Hu Y, et al. Clinical Characteristics of Coronavirus Disease 2019 in China [published online February 28, 2020]. N Engl J Med. DOI: 10.1056/NEJMoa2002032

[2] Novel Coronavirus Pneumonia Emergency Response Epidemiology Team. The epidemiological characteristics of an outbreak of 2019 novel coronavirus diseases (COVID-19) in China. Zhonghua Liu Xing Bing Xue Za Zhi. 2020, 41(2): 145–151.

[3] Wang C, Horby P W, Hayden F G, et al. A novel coronavirus outbreak of global health concern. Lancet. 2020, 395(10223): 470–473.

[4] Chan J F-W, Kok K-H, Zhu Z, et al. Genomic characterization of the 2019 novel human-pathogenic coronavirus isolated from a patient with atypical pneumonia after visiting Wuhan. Emerg Microbes Infect. 2020, 9(1): 221–236.

[5] WHO. Coronavirus disease (COVID-19) outbreak. 23 March 2020.. https://www.who.int/emergencies/diseases/novel-coronavirus-2019/situation-reports. Accessed March 23, 2020.

[6] Moher D, Liberati A, Tetzlaff J, et al. Preferred reporting items for systematic reviews and meta-analyses: the PRISMA statement. PLoS Med. 2009, 6(7): e1000097

[7] Stroup D F, Berlin J A, Morton S C, et al. Meta-analysis of observational studies in epidemiology: a proposal for reporting. Meta-analysis Of Observational Studies in Epidemiology (MOOSE) group. JAMA, 2000, 283(15): 2008–2012.

[8] Guidelines on diagnosis and treatment of novel coronavirus pneumonia (Trial sixth edition). Chinese Journal of Infection Control, 2020, 19(02): 192–195.

[9] Lau J, Ioannidis J P, Terrin N, et al. The case of the misleading funnel plot. BMJ, 2006, 333(7568): 597–600.

[10] Yuan Jing, S Y, Zuo Yujie, et al. Analysis of clinical characteristics of 223 patients with novel coronavirus pneumonia in Chongqing. Journal of Southwest University (Natural Science Edition). http://kns.cnki.net/kcms/detail/50.1189.N.20200305.1429.004.html. Accessed March 06, 2020.

[11] Wan Qiu, Shi Anqi, He Ting, Tang Lixin. Analysis of clinical features of 153 patients with novel coronavirus pneumonia in Chongqing. Chinese Journal of Clinical Infectious Diseases. DOI:10.3760/cma.j.cn115673-20200212-00030.

[12] Xiao Kaihu, Shui Lili, Pang Xiaohua, et al. The clinical features of the 143 patients with COVID-19 in North-East of Chongqing [published online February 27, 2020]. J Third Mil Med Univ. DOI:10.16016/j.1000-5404.202002097.

[13] Chen C, Chen C, Yan J T, et al. [Analysis of myocardial injury in patients with COVID-19 and association between concomitant cardiovascular diseases and severity of COVID-19]. Zhonghua xin xue guan bing za zhi, 2020, 48(0): E008.

[14] Chen G, Wu D, Guo W, et al. Clinical and immunologic features in severe and moderate forms of Coronavirus Disease 2019. medRxiv preprint. 2020: 2020.2002.2016.20023903.

[15] Jin-jin Zhang, Xiang Dong, Yi-Yuan Cao, et al. Clinical characteristics of 140 patients infected with SARS-CoV-2 in Wuhan, China [published online February 19, 2020]. Allergy. DOI:10.1111/all.14238.

[16] Liu Y, Sun W, Li J, et al. Clinical features and progression of acute respiratory distress syndrome in coronavirus disease 2019 [published online February 27, 2020]. medRxiv preprint. DOI:10.1101/2020.02.17.20024166.

[17] Liu J, Li S, Liu J, et al. Longitudinal characteristics of lymphocyte responses and cytokine profiles in the peripheral blood of SARS-CoV-2 infected patients [published online February 22, 2020]. medRxiv preprint. DOI: 10.1101/2020.02.16.20023671.

[18] Liu M, He P, Liu H G, et al. [Clinical characteristics of 30 medical workers infected with new coronavirus pneumonia]. Zhonghua Jie He He Hu Xi Za Zhi, 2020, 43(0): E016

[19] Tian S, Hu N, Lou J, et al. Characteristics of COVID-19 infection in Beijing. The Journal of infection. 2020: S0163-4453(0120)30101-30108.

[20] Liu J, Liu Y, Xiang P, et al. Neutrophil-to-Lymphocyte Ratio Predicts Severe Illness Patients with 2019 Novel Coronavirus in the Early Stage [published online February 12, 2020]. medRxiv preprint. DOI: 10.1101/2020.02.10.20021584.

[21] Cai Q, Huang D, Ou P, et al. COVID-19 in a Designated Infectious Diseases Hospital Outside Hubei Province,China [published online February 19, 2020]. medRxiv preprint. DOI:10.1101/2020.02.17.20024018.

[22] Xiaowei Fang, Qing Mei, Tianjun Yang, et al. Clinical characteristics and treatment strategies of 79 patients with COVID-19. Chinese Pharmacological Bulletin. http://kns.cnki.net/kcms/detail/34.1086.r.20200224.1340.002.html. Accessed February 25, 2020.

[23] Tianxin Xiang, Jiaming Liu, Fei Xu, et al. ZHANG Wei Analysis of clinical characteristics of 49 patients with Novel Coronavirus Pneumonia in Jiangxi province. Chinese Journal of Respiratory and Critical Care. http://kns.cnki.net/kcms/detail/51.1631.r.20200228.1506.002.html. Accessed March 02, 2020.

[24] Wan S, Yi Q, Fan S, et al. Characteristics of lymphocyte subsets and cytokines in peripheral blood of 123 hospitalized patients with 2019 novel coronavirus pneumonia (NCP) [published online February 12, 2020]. medRxiv preprint. DOI: 10.1101/2020.02.10.20021832.

[25] Juan Xiong, Wanli Jiang, Qian Zhou, Xiaoqing Hu, Chunying Liu. Clinical characteristics, treatment, and prognosis in 89 cases of COVID-2019 [published online March 03, 2020]. Wuhan University Journal (Medical Edition). DOI: 10.14188/j.16718852.2020.0103

[26] Zhihui Dai, Li dong Gao, Kai wei Luo, et al. Clinical characteristics analysis of novel coronavirus pneumonia in Hunan province. Practical Preventive Medicine. http://kns.cnki.net/kcms/detail/43.1223.R.20200305.1537.005.html. Accessed March 06, 2020.

[27] Chen Lei, Liu Huiguo, Liu Wei, et al. Analysis of clinical features of 29 patients with 2019 novel coronavirus pneumonia. Zhonghua Jie He He Hu Xi Za Zhi, 43(3), 203–208. DOI:10.3760/cma.j.issn.1001-0939.2020.03.013.

[28] Wen Ke, Li Wengang, Zhang Dawei, et al. Epidemiological and clinical characteristics of 46 newly-admitted coronavirus disease 2019 cases in Beijing. DOI:10.3760/cma.j.cn311365-20200219-00086

[29] Huang C, Wang Y, Li X, et al. Clinical features of patients infected with 2019 novel coronavirus in Wuhan, China. Lancet, 395(10223), 497–506.

[30] Chen N, Zhou M, Dong X, et al. Epidemiological and clinical characteristics of 99 cases of 2019 novel coronavirus pneumonia in Wuhan, China: a descriptive study. Lancet, 395(10223), 507–513.

[31] Zhu N, Zhang D, Wang W, et al. A Novel Coronavirus from Patients with Pneumonia in China, 2019. N Engl J Med. 2020, 382(8): 727–733.

[32] Yang Sijue G H. Use Stones from Another Mountain to Polish One’s Jade: Learn from MERS Studies toexplore potential mechanisms underlying the effect of diabetes mellitus on COVID-19. Chin J Endocrinol Metab, 2020, (00): E001–E001

[33] Zhao Y, Zhao Z, Wang Y, et al. Single-cell RNA expression profiling of ACE2, the putative receptor of Wuhan 2019-nCov [published online January 26, 2020]. bioRxiv preprint. DOI:10.1101/2020.01.26.919985

[34] Jia X, Yin C, Lu S, et al. Two Things About COVID-19 Might Need Attention [published online January 23, 2020]. Preprints. DOI:10.20944/preprints202002.0315.v1.

[35] Yang X, Yu Y, Xu J, et al. Clinical course and outcomes of critically ill patients with SARS-CoV-2 pneumonia in Wuhan, China: a single-centered, retrospective, observational study. Lancet Respir Med, 2020: S2213-2600(2220)30079-30075.

[36] Gu J, Gong E, Zhang B, et al. Multiple organ infection and the pathogenesis of SARS. J Exp Med, 2005, 202(3): 415–424.

[37] Chen Y, Guo D. Molecular mechanisms of coronavirus RNA capping and methylation[J]. Virol Sin, 2016, 31(1): 3–11.

[38] Wang L F, Shi Z, Zhang S, et al. Review of bats and SARS. Emerg Infect Dis, 2006, 12(12): 1834–1840.

[39] Lu R, Zhao X, Li J, et al. Genomic characterisation and epidemiology of 2019 novel coronavirus: implications for virus origins and receptor binding. Lancet. 2020, 395(10224): 565–574.

[40] Expert Group on Prevention and Control of COVID-19 of Chinese Preventive Medicine Association. [An update on the epidemiological characteristics of novel coronavirus pneumonia(COVID-19)]. Chinese Journal of Virology. DOI:10.16505/j.2095-0136.2020.0015

[41] Luo W,Yu H,Gou J,Li X,Sun Y,Li J,Liu L. Clinical Pathology of Critical Patient with Novel Coronavirus Pneumonia (COVID-19). Preprints. 2020, 2020020407.

[42] H S, X H, N J, et al. Radiological findings from 81 patients with COVID-19 pneumonia in Wuhan, China: a descriptive study [published online February 24, 2020]. Lancet Infect Dis. DOI: 10.1016/S1473-3099(20)30086-4.

[43] He L, Ding Y-Q, Che X-Y, et al. Expression of the monoclonal antibody against nucleocapsid antigen of SARS-associated coronavirus in autopsy tissues from SARS patients. Di 1 jun yi da xue xue bao = Academic journal of the first medical college of PLA. 2003, 23(11): 1128–1130.

[44] Zou Xin,Chen Ke,Zou Jiawei et al. Single-cell RNA-seq data analysis on the receptor ACE2 expression reveals the potential risk of different human organs vulnerable to 2019-nCoV infection [published online March 12, 2020]. Front Med. DOI: 10.1007/s11684-020-0754-0.

